# von Willebrand factor-related values can predict bleeding events in patients with left ventricular assist devices

**DOI:** 10.1101/2023.12.15.23299936

**Authors:** Ryuichi Taketomi, Ko Sakatsume, Shintaro Katahira, Kota Goto, Misako Suzuki, Zuo Yunan, Konosuke Sasaki, Midori Miyatake, Katsuhiro Hosoyama, Koki Ito, Yusuke Suzuki, Goro Takahashi, Kiichiro Kumagai, Hisanori Horiuchi, Yoshikatsu Saiki

## Abstract

**Background:** Recent advances in left ventricular assist device (LVAD) therapy have significantly contributed to the improved management of severe heart failure. The unignorable risk of bleeding events associated with the device therapy, however, remains to be addressed. Predictive factors for bleeding events are not fully defined.

**Methods:** Patients implanted with various types of LVADs were assessed for von Willebrand factor (VWF)-related values and platelet proteins from 2011 to 2023 at Tohoku University Hospital. We evaluated the relationship between these parameters and bleeding events at remote periods after LVAD implantation.

**Results:** The VWF large multimer index (VWF-LMI), the ratio of VWF ristocetin cofactor activity to VWF antigen level (VWF:RCo/VWF:Ag), and maximal ristocetin (1.2 µg/mL) induced platelet aggregation (RIPA) rates decreased in a pump speed- and device type-dependent manner. Major bleeding events, defined according to the Interagency Registry for Mechanically Assisted Circulatory Support criteria, occurred in 28.8% (19/66) of the patients. Kaplan–Meier methods, log-rank tests, and Cox regression analyses revealed that VWF-LMI <42.3 (p=0.0002), VWF:RCo/VWF:Ag <0.52 (p=0.0239), and maximal RIPA rate <79.2 (p=0.0012) predicted bleeding events with hazard ratios of 6.96, 2.89, and 13.4, respectively. The platelets of the patients with LVAD support exhibited significantly reduced expression levels of glycoprotein (GPVI; p=0.0334) and glycoprotein b (GPIb; p=0.0061) compared with healthy subjects. Nevertheless, patients with bleeding events did not exhibit reduced GPVI (p=0.5267) or GPIb (p=0.6674) levels compared to those without bleeding events.

**Conclusions:** VWF-related values, including VWF-LMI, VWF:RCo/VWF:Ag, and maximal RIPA rate, but not reduced platelet proteins, predicted bleeding events in patients with LVADs.

**Registration:** URL: https://www.umin.ac.jp/ctr; Unique identifier: UMIN000027761

**Clinical Perspective:** *What Is New?:*

- Comprehensive measurements of von Willebrand factor-related values, platelet proteins, and ristocetin-induced platelet aggregation rates were obtained in patients with left ventricular assist device support to evaluate their association with bleeding events.
- The von Willebrand factor large multimer index, ratio of von Willebrand factor ristocetin cofactor activity to von Willebrand factor antigen levels, and maximal ristocetin-induced platelet aggregation rate predicted bleeding events in patients implanted with a left ventricular assist device.

*What Are the Clinical Implications?:*

- Predictors of bleeding events in patients with left ventricular assist device support were identified in this study. Understanding how von Willebrand factor-related values are associated with bleeding events will facilitate the management of potential bleeding events.
- The ratio of von Willebrand factor ristocetin cofactor activity to von Willebrand factor antigen levels is a predictor of bleeding events that can be measured and evaluated relatively easily in many centers without special techniques or equipment, in contrast to analysis of the von Willebrand factor large multimer index.

## Introduction

Left ventricular assist devices (LVADs) have made tremendous contributions to the treatment of severe heart failure; however, they are associated with complications such as infection, thrombosis, stroke, and bleeding events, which is most commonly gastrointestinal tract bleeding ^1–3^.

LVAD-associated bleeding is thought to be caused by unphysiologically high shear stress inside the pump, which causes degradation of high-molecular-weight (large) von Willebrand factor (VWF) multimers having a critical role in hemostasis, leading to a hemostatic disorder known as acquired von Willebrand syndrome (AVWS) ^4,5^. Platelet function is also reportedly impaired in patients with LVADs due to the shedding of glycoprotein (GPVI) and glycoprotein b (GPIb), platelet proteins with important roles in hemostasis ^6–9^. Several months after LVAD implantation, fragile and abnormal vessels called angiodysplasia lesions develop in the gastrointestinal tract just underneath the epithelial cells ^10–12^. Thus, gastrointestinal bleeding at remote periods after implantation typically derives from angiodysplasia lesions under impaired hemostatic conditions due to dysfunction of VWFs and/or platelets.

Recently developed LVADs with a lower shear stress in the pump seem to be related with fewer LVAD-associated gastrointestinal bleeding events^13,14^. Nevertheless, bleeding events remain a common complication in patients with LVADs, accounting for nearly 20.0% of hospital readmissions ^15^. Patients with LVADs exhibit a remarkably severe loss of VWF large multimers compared to patients with severe aortic stenosis based on the VWF large multimer index (VWF-LMI), a quantitative score we proposed ^5,16–24^ and have widely used ^25–27^ to evaluate VWF large multimers in VWF multimer analysis. In the present study, we demonstrated that patients with a bleeding event had significantly lower VWF-LMI values compared to patients without a bleeding event ^18^.

The strength of the contribution of VWF impairment and/or platelet dysfunction to bleeding events in patients with LVADs remains unclear. More importantly, no common measures to predict LVAD-associated bleeding events have been developed. To address these issues, we evaluated the association between bleeding events in patients with LVADs and their VWF-related values, including VWF-LMI and platelet protein levels, in patients who underwent LVAD implantation at Tohoku University Hospital.

## Methods

### Participants

Patients who underwent LVAD implantation at Tohoku University Hospital between July 2011 and June 2023, were analyzed. Hematologic analyses were performed when patients were stable. The association between the results of the hematologic analyses and “major bleeding” events as defined in the IMACS criteria, including suspected internal or external bleeding resulting in death, re-operation, hospitalization, and/or transfusion of red blood cells^28^, excluding perioperative events, was evaluated.

We previously reported an analysis of 41 patients who underwent LVAD implantation and were followed at Tohoku University^18^. Of those 41 patients, 2 patients were excluded from the analysis as the analyzed blood samples were not obtained under a stable condition (Supplementary figure 1) and 39 patients were analyzed here. Following that report, another 33 patients underwent LVAD implantation at Tohoku University Hospital by June 2023. Excluding 6 patients who declined to participate, transferred to another hospital, or died from postoperative complications, 27 patients were enrolled in the present study. Thus, a total 66 patients with LVADs were analyzed here.

Platelet proteins and ristocetin-induced platelet aggregation (RIPA) levels were analyzed in 30 of the 66 patients implanted with LVADs by September 2022, and in 10 enrolled healthy subjects who were not currently taking any medications.

### Laboratory data

Blood samples, collected in sodium citrate at least 1 month after LVAD implantation when patients were stable, were centrifuged at 150 × g for 10 min at room temperature to isolate platelet-rich plasma (PRP). Centrifugation at 2,000 × g for 5 min at room temperature was performed to obtain platelet poor plasma (PPP). Within 2 h after obtaining the PRP and PPP samples, 1.2 mg/mL ristocetin was added and RIPA was measured using an automated blood coagulation analyzer (CN6000, Sysmex Corp, Kobe, Japan) according to the manufacturer’s instructions. The PRP was also utilized for platelet isolation through gel filtration chromatography ^29^. Isolated platelets were suspended in Laemmli’s sodium dodecyl sulfate-containing buffer with 2-mercaptoethanol and stored at -80°C until use. In parallel, the collected blood samples were centrifuged at 1,200 × g for 10 min at 4°C to obtain plasma. The obtained plasma was stored at -80°C until use.

The VWF ristocetin cofactor activity (VWF:RCo) and VWF antigen levels (VWF:Ag) in the plasma samples from each of the patients were measured with the CN6000 (Sysmex Corp) according to the manufacturer’s instructions.

### von Willebrand factor large multimer index

VWF-LMI was analyzed using a 1.0% agarose gel to which equal amounts of VWF:Ag were applied under non-reducing conditions followed by Western blotting using FITC-labelled polyclonal rabbit anti-human VWF antibody (DAKO, Glostrup, Denmark) as the primary antibody. The Western blotting analysis probing VWF was visualized by ImmunoStarVR Zeta chemiluminescence (Wako, Osaka, Japan) and evaluated using Amersham ImageQuant800 (Cytiva, Marlborough, MA, USA) and ImageJ. Bands above the 10^th^ lowest band were classified as large multimers. Based on densitometric analysis, the VWF-LMI was calculated as the percentage of a patient’s VWF large multimer ratio to that of the control (Siemens Standard plasma) analyzed in the lane adjacent to the patient’s plasma, as described previously^5,16–23,25–27^.

### Western blotting for platelet proteins evaluation

Proteins in the platelet lysate of each patient and control subject, which contained an equal amount of tubulin, were separated by sodium-dodecyl sulphate polyacrylamide-gel electrophoresis using 12.5% e-PAGEL gel (ATTO, Tokyo, Japan). They were then transferred to a nitrocellulose membrane, Protran BA85 (Cytiva), and Western blotting was performed. The primary antibodies used in this study are listed in Supplementary figure 2. Secondary antibodies were mouse anti-human IgG (Jackson ImmunoResearch, West Grove, PA, USA), rabbit anti-human IgG (Jackson ImmunoResearch) or sheep anti-human IgG (R&D Systems, MN, USA). It was finally visualized by ImmunoStar Zeta chemiluminescence (Wako) and quantified using Amersham ImageQuant800 (Cytiva) and ImageJ, using the amount of a specific healthy person as 100%.

### Statistical analyses

All statistical analyses were performed using GraphPad Prism version 10 (GraphPad Software, San Diego, CA, USA). Continuous variables are expressed as mean ± standard deviation. Categorical variables are presented as frequencies and percentages, and inter-group comparisons of categorical variables were analyzed using Fisher’s exact test or the χ2 test. Intergroup comparisons of continuous variables were performed using Student’s t-test or the Mann Whitney test if normally distributed. Bleeding event-free survivals were analyzed using the Kaplan–Meier method, log-rank test, and Cox regression analysis. Differences were considered significant when the P-value was less than 0.050.

### Ethics

This study was performed in accordance with the Declaration of Helsinki. The study protocol was approved by the Institutional Ethics Committee in Tohoku University (Certification number; 2017-1-386 and 2020-1-935). The study protocol was also registered in the University Hospital Medical Information Network (UMIN) (UMIN number; UMIN000027761). Written informed consent was obtained from each participant.

## Results

### Participant characteristics

In this study, we analyzed 66 patients with LVADs, as described above (Supplementary figure 1). Age, sex, and blood type did not differ significantly between healthy subjects and patients (Table 1). Patients underwent implantation of various types of LVADs; a centrifugal-type LVAD, including the EVAHEART (Sun Medical Technology Research Corp., Nagano, Japan; n=9), HeartMate3 (Abbott, Chicago, IL, USA; n=12), HVAD (Medtronic, Framingham, MA, USA; n=4), DuraHeart (Terumo, Tokyo, Japan; n=4); or an axial-type LVAD, including the Jarvik2000 (Jarvik Heart Inc, New York, NY, USA; n=10) and HeartMate (Abbott, Chicago, IL, USA; n=27;Table 1).

**Table 1.**
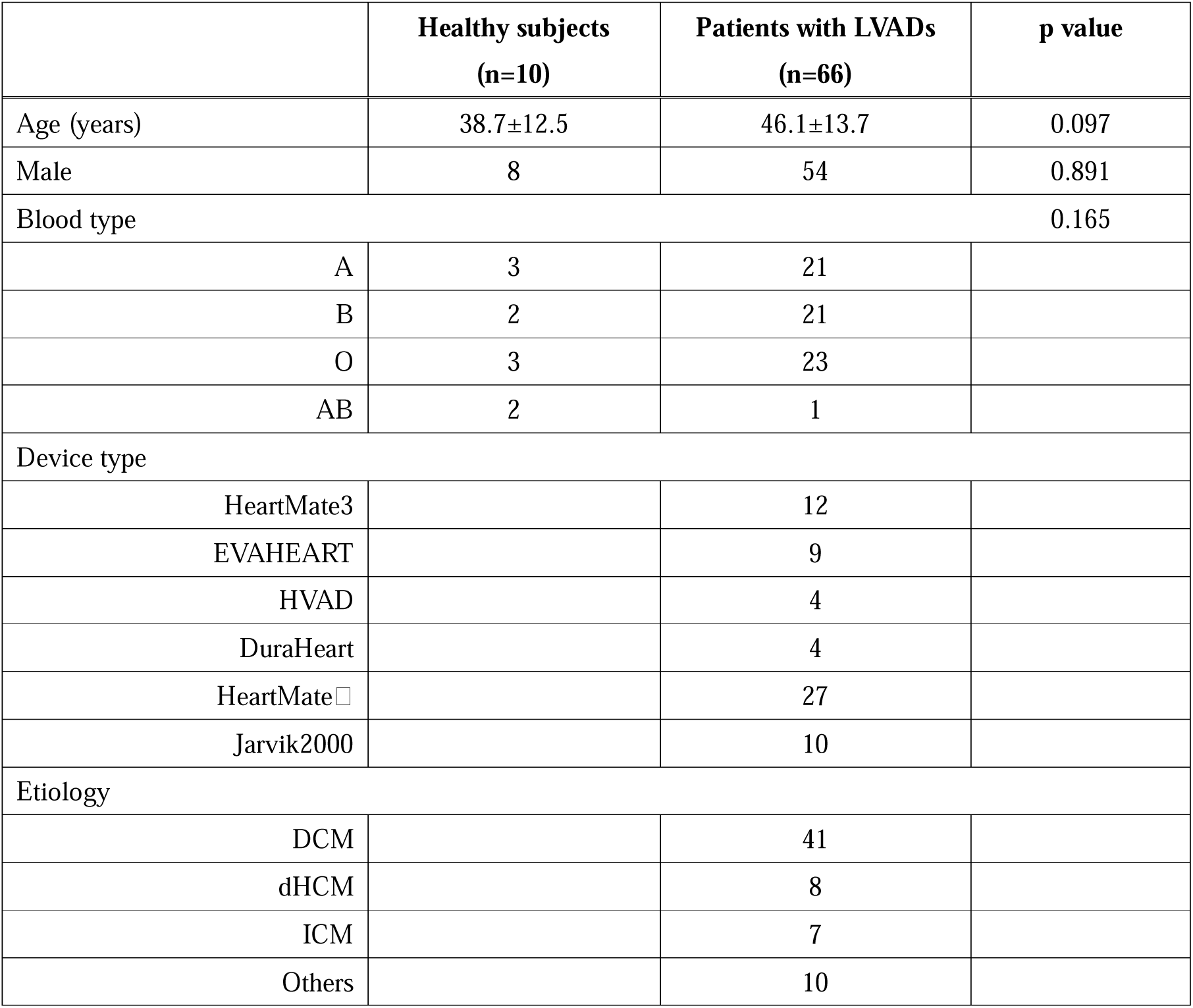
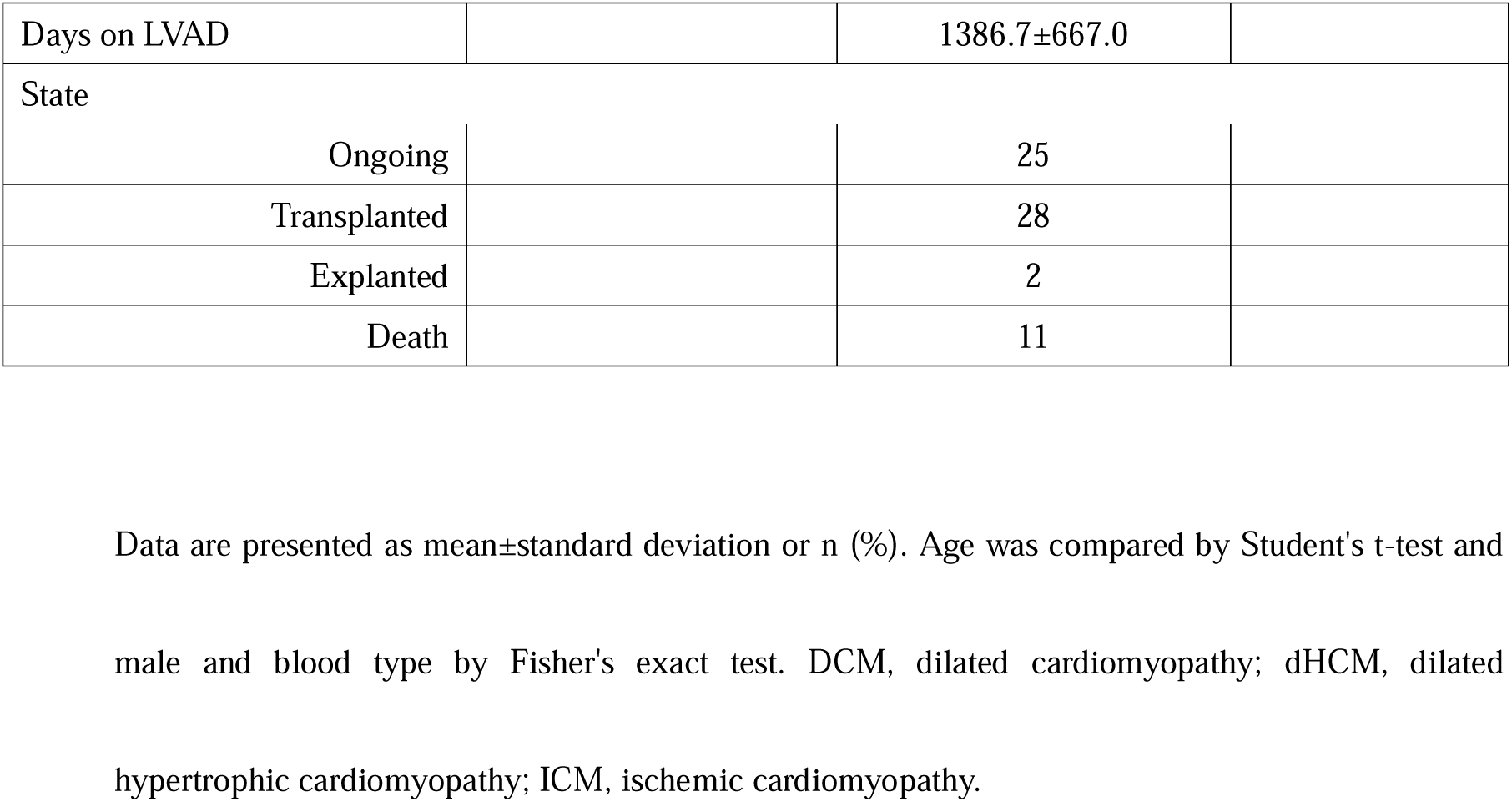
Characteristics of participants in this study.

### Evaluation of VWF-LMI

VWF multimer analysis of the patients’ plasmas was performed by Western blotting under non-reducing conditions (Figure 1A). VWF large multimers were evaluated with the VWF-LMI as described in the Methods. The mean VWF-LMI of the patients was 47.9 ± 25.4%. We have proposed that VWF-LMI<80.0% reasonably indicates loss of VWF large multimers^24^, and 57 of 66 patients (86.3%) had VWF-LMI<80.0%. Patients with a centrifugal-type LVAD had a higher VWF-LMI than patients with an axial-type LVAD (p<0.001), and patients implanted with an HVAD and DuraHeart had a lower VWF-LMI (Figure 1B). The low VWF-LMIs in patients implanted with an HVAD and DuraHeart having a low pump speed might be due to their outlets with narrow diameters. The VWF-LMI decreased in a pump speed-dependent manner (p<0.001, r=-0.599; Figure 1C).

**Figure 1.** (A) Distribution of von Willebrand factor (VWF) large multimers. Bands above the 10^th^ lowest were classified as large multimers. Compared with VWF large multimers in control subjects, VWF large multimers of patients with the HeartMate3 were reduced and those of patients with the Jarvik2000 were even further reduced. (B) The VWF large multimer index (VWF-LMI) differed significantly between patients implanted with the EVAHEART and HeartMate (p<0.001), EVAHEART and Jarvik2000 (p<0.001), HeartMate3 and HeartMate (p<0.001), and the HeartMate3 and Jarvik2000 (p<0.001). (C) VWF-LMI decreased in a pump speed-dependent manner (p<0.001, r=-0.599).

### Evaluation of VWF:RCo/VWF:Ag

The ratio of VWF:RCo to VWF:Ag can be measured in plasma by an automated blood coagulation analyzer. VWF:RCo/VWF:Ag <0.70 is used for the diagnosis of hereditary von Willebrand disease. The mean VWF:RCo/VWF:Ag for the 66 patients with LVADs was rather low (0.53±0.22), and 51 of 66 patients (77.3%) had VWF:RCo/VWF:Ag <0.70. The VWF-LMI positively correlated with the VWF:RCo/VWF:Ag (p<0.001, r=0.468; Figure 2A). Nevertheless, VWF:RCo/VWF:Ag did not correlate with the device type or pump speed (Figure 2B and C).

**Figure 2.** (A) The VWF large multimer index (VWF-LMI) positively correlated with the ratio of VWF ristocetin cofactor activity to VWF antigen levels (VWF:RCo/VWF:Ag; p<0.001, r=-0.468). (B) Association between device type and VWF:RCo/VWF:Ag. No significant difference was detected between device types. (C) Pump speed did not correlate with VWF:RCo/VWF:Ag (p=0.170, r=-0.171).

### Evaluation of RIPA

Ristocetin is known to induce GPIb platelet protein binding to VWF, which causes platelet aggregation. We analyzed RIPA in 30 patients with LVADs among the enrolled 66 patients as shown in Supplementary figure 1. The mean maximal RIPA rates were 81.4±11.9%, and did not differ significantly from those of healthy subjects, 82.0±8.3% (p=0.834). Nevertheless, VWF-LMI positively correlated with the maximal RIPA rate (p<0.001, r=0.660; Figure 3A). Patients with a centrifugal-type LVAD had a higher maximum RIPA rate compared to patients with an axial-type LVAD (p=0.004) and these values differed significantly between the EVAHEART and Jarvik2000 when comparing each device individually (p=0.010; Figure 3B). Maximal RIPA rates decreased in a pump-speed dependent manner (p<0.001, r=-0.664; Figure 3C).

**Figure 3.** (A) The VWF large multimer index (VWF-LMI) positively correlated with maximal ristocetin-induced platelet aggregation (RIPA) rate (p<0.001, r=-0.660). (B) Association between device type and maximal RIPA rate. The association of the maximal RIPA rate differed significantly between the EVAHEART and Jarvik 2000 (p=0.001). (C) Maximal RIPA rates decreased in a pump speed-dependent manner (p<0.001 r=-0.664).

### Bleeding events

Nineteen patients developed bleeding events among the enrolled 66 patients during the observation period until heart transplantation, death, or bleeding event. Patients developing a bleeding event are summarized in Table 2. Of the 19 patients who developed a bleeding event, 13 had gastrointestinal bleeding. Cases 4, 7, 8, and 14 were admitted for evaluation of anemia despite having no bleeding symptoms. All patients were kept on warfarin. Their prothrombin time-international normalized ratios (PT-INRs) were within or below the target range (EVAHEART: 2.5-3.5, other devices: 2.0-3.0) except 1 patient whose PT-INR exceeded only slightly the target range at 3.13. Comparison of patients with bleeding events (Bleeding (+)) and those without bleeding events (Bleeding (-)) by univariate analysis revealed significant differences in device type, days on LVAD, VWF-LMI, VWF:RCo/VWF:Ag, lactate dehydrogenase, and platelet counts (Table 3). Following multivariate analysis with these factors, however, only VWF-LMI remained statistically significant (p=0.045).

**Table 2.** List of patients developing bleeding events.

| Case | Days on LVAD | Sex | Symptoms | Bleeding site | Device type | Antiplatelet agents | Hb (g/dL) | PT-INR | Plt ($10^3/\mu\text{L}$) |
| --- | --- | --- | --- | --- | --- | --- | --- | --- | --- |
| 1 | 17 | Male | black stool | intestine | Jarvik2000 | aspirin 100 mg | 6.9 | 1.79 | 20.2 |
| 2 | 32 | Male | melena | ascending colon | Jarvik2000 | aspirin 100 mg | 7.7 | 3.13 | 11.8 |
| 3 | 142 | Male | black stool | intestine | HeartMate□ | aspirin 100 mg | 7.8 | 2.85 | 18.4 |
| 4 | 159 | Male | no symptoms | unknown | HeartMate□ | aspirin 100 mg | 6.3 | 2.23 | 32.4 |
| 5 | 209 | Male | black stool | stomach | HeartMate□ | aspirin 100 mg | 7.2 | 2.00 | 13.5 |
| 6 | 266 | Male | black stool | stomach | Jarvik2000 | aspirin 100 mg | 7.1 | 1.90 | 14.5 |
| 7 | 311 | Male | no symptoms | unknown | HeartMate3 | aspirin 100 mg | 7.2 | 1.96 | 23.8 |
| 8 | 609 | Male | no symptoms | unknown | HVAD | clopidogrel 75 mg | 8.5 | 1.52 | 14.6 |
| 9 | 674 | Male | black stool | intestine | HeartMate□ | aspirin 100 mg | 8.5 | 2.35 | 25.3 |
| 10 | 685 | Male | melena | ascending colon | Jarvik2000 | aspirin 100 mg + clopidogrel 75 mg | 6.6 | 2.27 | 19.1 |
| 11 | 791 | Male | black stool | intestine | HeartMate□ | aspirin 100 mg | 7.7 | 2.65 | 23.0 |
| 12 | 833 | Male | black stool | ascending colon | Jarvik2000 | aspirin 100 mg + clopidogrel 75 mg | 12.4 | 2.84 | 9.4 |
| 13 | 843 | Male | melena | ascending colon | Jarvik2000 | clopidogrel 75 mg | 7.4 | 2.40 | 32.2 |
| 14 | 868 | Male | no symptoms | unknown | HVAD | aspirin 100 mg | 7.1 | 1.18 | 19.4 |
| 15 | 1064 | Female | excessive menstruation | uterus | HeartMate□ | aspirin 100 mg | 7.3 | 2.81 | 20.8 |
| 16 | 1110 | Male | melena | ascending colon | HeartMate□ | aspirin 100 mg | 6.1 | 2.73 | 27.1 |
| 17 | 1123 | Male | black stool | intestine | HeartMate ☐ | aspirin 100 mg | 9.1 | 2.29 | 20.9 |
| 18 | 1627 | Female | excessive<br>menstruation | ovary | Jarvik2000 | clopidogrel 50 mg | 6.5 | 1.77 | 14.7 |
| 19 | 2470 | Female | melena | intestine | Jarvik2000 | aspirin 100mg | 5.4 | 1.43 | 13.1 |

Bleeding events defined by the Interagency Registry for Mechanically Assisted Circulatory Support criteria including suspected internal or external bleeding, resulting in death, re-operation, hospitalization, and/or transfusion of red blood cells<sup>28</sup>, excluding perioperative events<sup>28</sup>. Hb, hemoglobin; LVAD, left ventricular assist device; PT-INR, prothrombin time-international normalized ratio; Plt, platelet count.

**Table 3.**
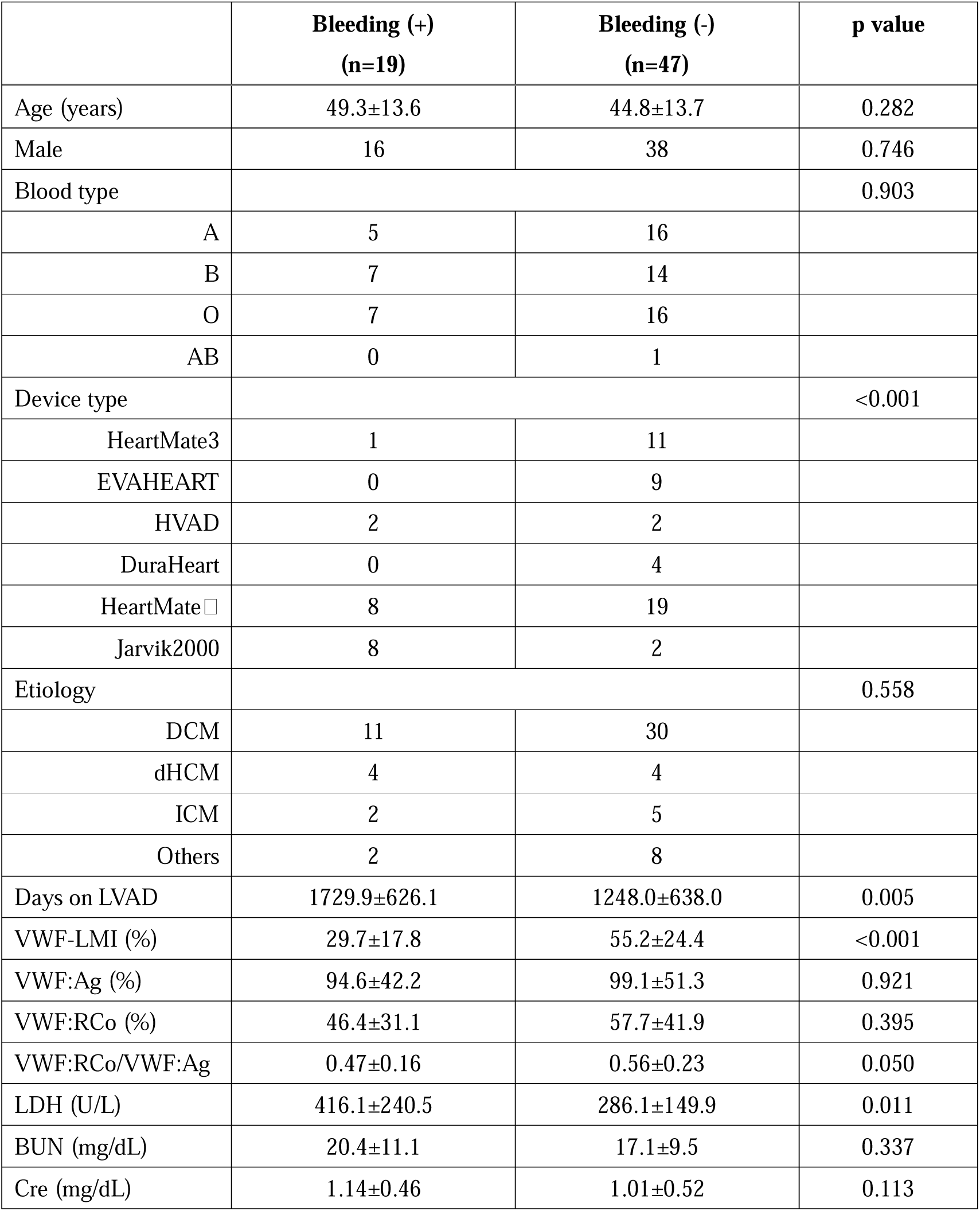
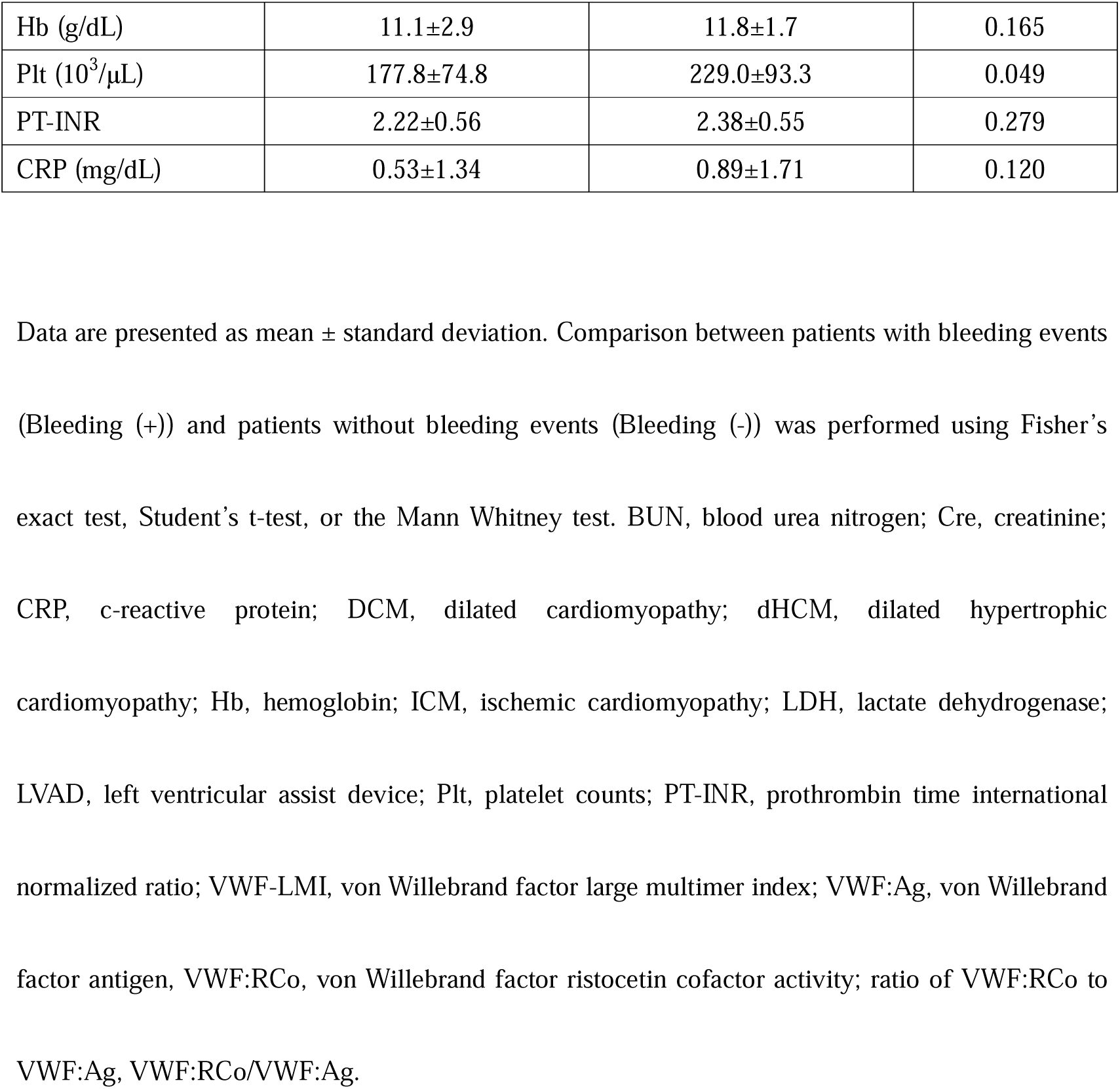
Comparison of characteristics and laboratory data between patients with bleeding events (Bleeding (+)) and patients without bleeding events (Bleeding (-))

### Kaplan-Meier curves for bleeding events

The receiver operating characteristics (ROC) analysis determined a cutoff value of 42.3% for VWF-LMI in relation to bleeding events (sensitivity 0.68, specificity 0.84). Using this cutoff value, patients were divided into 2 groups and the Kaplan-Meier curve revealed a significant increase in bleeding events in patients with VWF-LMI <42.3% compared to patients with VWF-LMI ≧42.3% (p<0.001, hazard ratio [HR] 6.96 [95% confidence interval [CI] 2.81-17.2]; Figure 4A). The ROC analysis determined a cutoff value of 0.52 for VWF:RCo/VWF:Ag with respect to bleeding events (sensitivity 0.68, specificity 0.68). Using this cutoff value, patients were divided into 2 groups and the Kaplan-Meier curve revealed a significant increase in bleeding events in patients with VWF:RCo/VWF:Ag <0.52 compared to patients with VWF:RCo/VWF:Ag ≧0.52 (p=0.024, HR 2.89 [95%CI 1.17-7.17]; Figure 4B)

**Figure 4.** Comparison between patients in the 2 groups based on cut-off values determined by receiver operating characteristics curve, using Kaplan-Meier method, log-rank test, and Cox regression analysis revealed (A) a significant increase in bleeding events in the patients with a VWF large multimer index (VWF-LMI) <42.3 compared to the patients with VWF-LMI ≧42.3 (p<0.001, HR 6.96), (B) a significant increase in bleeding events in the patients with a ratio of VWF ristocetin cofactor activity to VWF antigen levels (VWF:RCo/VWF:Ag) <0.52 compared to patients with VWF:RCo/VWF:Ag ≧0.52 (p=0.024, HR 2.89), and (C) a significant increase in bleeding events in patients with maximal ristocetin-induced platelet aggregation (RIPA) rates <79.2 compared to patients with maximal RIPA rates ≧ 79.2 (p=0.001, HR 13.4). VWF-LMI <42.3, VWF:RCo/VWF:Ag <0.52, and maximal RIPA rate <79.2 predicted bleeding events.

Six of the 30 patients (20.0%) analyzed for RIPA developed bleeding events. The ROC analysis determined a cutoff value of 79.2% for maximal RIPA rate with respect to bleeding events (sensitivity 0.79, specificity 0.83). Using this cutoff value, patients were divided into 2 groups and the Kaplan-Meier curve revealed a significant increase in bleeding events in the patients with a maximal RIPA rate <79.2% compared to patients with a maximal RIPA rate ≧79.2% (p=0.001, HR 13.4 [95%CI 2.22-81.17]; Figure 4C)

### Evaluation of platelet proteins

We first confirmed the high specificity of each primary antibody to ensure its adequacy for use in the present study (Figure 5A). With the antibodies, platelets isolated from 30 patients were analyzed by Western blotting by calibrating the PRP volumes based on the amount of tubulin in a control healthy subject. Each result was expressed as a percentage of the control healthy subject’s expression level. Among 11 platelet proteins, including PAR-1, GPIIIa, P2Y12, and P-selectin, lower expression levels of GPIb (89.3±6.3 vs 77.4±18.2, p=0.006) and GPVI (108.8±9.0 vs 81.3±33.6, p=0.033) were observed in patients with LVADs (n=30) compared to healthy subjects (n=10). Expression levels of PAR-1 (p=0.210), TXA2R (p=0.221), GPIIIa (p=0.842), FcRγ (p=0.678), P2Y1 (p=0.072), P2Y12 (p=0.188), 5-HTR2 (p=0.286), PGI2R (p=0.770), and P-selectin (p=0.358) did not differ significantly between patients and controls (Figure 5B). The levels of GPIb or GPVI were not associated with the device type or pump speed. Further, the levels of GPIb and GPVI were not different between Bleeding (+) and Bleeding (-) (p=0.667 and p=0.890, respectively) (Supplementary figure 3).

**Figure 5.** (A) Western blotting of each primary antibody. Western blotting was performed on 11 platelet proteins involved in platelet aggregation, including PAR-1, TXA2R, GPIb, GP IIIa, GP VI, FcRγ, P2Y1, P2Y12, 5HTR-2, PGI2R and P-selectin. (B) Comparison of the levels of platelet proteins, including (a) PAR-1, (b) TXA2R, (c) GP b, (d) GP a, (e) GP, (f) FcRγ, (g) P2Y1, (h) P2Y12, (i) 5HTR-2, (j) PGI2R and (k) P-selectin, between healthy subjects and patients with left ventricular assist devices (LVADs). Of the 11 platelet proteins, only GPIb (p=0.006) and GPⅥ (p=0.033) were significantly reduced in patients with LVADs. No significant differences in PAR-1 (p=0.210), TXA2R (p=0.221), GPIIIa (p=0.842), FcRγ(p=0.678), P2Y1 (p=0.072), P2Y12 (p=0.188), 5HTR-2 (p=0.286), PGI2R (p=0.770), and P-selectin (p=0.358) were detected between healthy subjects and patients with LVADs. Levels of platelet proteins were not associated with bleeding events.

## Discussion

The findings of the present study demonstrated that 19 (28.8%) of the 66 patients with LVADs developed bleeding events during a mean follow-up of 1386.7±667.0 days after LVAD implantation. Patients with bleeding events (n=19) had lower VWF-LMI, VWF:RCo/VWF:Ag, and maximal RIPA rate than those without bleeding events. The Kaplan-Meier curve showed that VWF-LMI <42.3% (p<0.001), VWF:RCo/VWF:Ag <0.52 (p=0.024), and maximal RIPA rate <79.2% (p=0.001) predicted bleeding events with HRs of 6.96, 2.89, and 13.42, respectively. Thus, we have first found predictive factors for bleeding events in patients with LVAD that are VWF-related values including VWF-LMI, VWF:RCo/VWF:Ag and maximal RIPA rates.

VWF:RCo/VWF:Ag <0.70 is used to diagnose von Willebrand disease type 2A ^30^ and can be measured by an automated hemostatic analyzer in ordinary hospitals. Thus, measuring this ratio is much more feasible than measuring VWF-LMI that requires a special Western blotting technique for analyzing huge VWF multimers of ∼20,000,000 daltons, and RIPA that requires freshly collected patient’s platelets and a light-transmission platelet aggregometer. Furthermore, we have recently reported that VWF:RCo/VWF:Ag <0.70 predicts a loss of VWF large multimers, defined as VWF-LMI <80.0%, with high specificity, but low sensitivity in patients with severe aortic stenosis (AS)^24^. This is probably due to the relatively low degree of loss of VWF large multimers in AS patients having VWF-LMI of 70.0%-80.0% and many AS patients exhibited VWF:RCo/VWF:Ag around 0.70^24^. VWF:RCo/VWF:Ag in many AS patients were overlapped with those in healthy subjects. Therefore, use of VWF:RCo/VWF:Ag could be limited for the diagnosis of AVWS in AS patients. In contrast, VWF-LMIs in patients with LVADs in the present study were 20.0%-80.0% (47.9 ± 25.4%), which were much lower compared to those in patients with severe AS. Accordingly, the positive predictive value of VWF:RCo/VWF:Ag < 0.70 for VWF-LMI < 80.0% is very high with 45/51 (88.2%). Thus, VWF:RCo/VWF:Ag <0.70 could be a useful indicator to monitor the loss of VWF large multimers in patients with LVADs.

The VWF:RCo/VWF:Ag correlated with VWF-LMI. The majority (51/66; 77.3%) of patients had a VWF:RCo/VWF:Ag <0.70. Importantly, VWF:RCo/VWF:Ag <0.52 predicted bleeding events (p=0.024) with an HR of 2.89 as described above. Thus, VWF:RCo/VWF:Ag may be a useful predictor in centers where VWF-LMI and RIPA are difficult to measure. Since VWF-LMI, which predicted bleeding events with an HR 6.96, is a better predictor than VWF:RCo/VWF:Ag, VWF-LMI is recommended in centers where it can be measured.

Maximal RIPA rates also predicted bleeding events, as did VWF-LMI and VWF:RCo/VWF:Ag. However, the reduction in maximal RIPA rates was smaller, with a mean maximal RIPA rates of 81.4% and a cut-off value of 79.2% for bleeding events in 30 patients with LVAD. This may be due to high ristocetin concentrations used for the purpose of differentiating von Willebrand disease.

We recently demonstrated individual variability in VWF fragility in response to shear stress ex vivo^21^. In this study, VWF-LMI varied from 40.0%-90.0%, even in patients with the HeartMate3, which is currently the most frequently implanted LVAD in the world and is associated with fewer bleeding events, indicating individual differences in VWF vulnerability to shear stress^14^. It is therefore important to measure VWF-related factors, since even in patients with LVADs producing low shear stress, there are patients with a degraded VWF large multimer.

Regarding the association between bleeding events and the expression levels of platelet proteins, Mondal et al reported that patients with LVADs (n=51) had lower expression levels of GPIb and GPVI than healthy subjects, and that patients with bleeding events had lower GPIb and GPVI expression levels than those without bleeding events^9^. It is unclear why decreased GPIb or GPVI was not associated with bleeding events in our study. We consider following reasons; (1) Mondal et al analyzed patients treated with HeartMateII, Jarvik2000, and HVAD^9^, which degraded VWF multimers more severely shown in this study. They did not, however, analyze VWF. It is highly likely that the patients with bleeding events in their report had reduced VWF large multimers as well as reduced levels of platelet proteins. (2) Although the definitions of bleedings were based on the INTERMACS criteria in both studies, our study excluded perioperative events due to the greater influence of the surgical technique and unstable hemodynamics in the immediate postoperative period. It is possible that platelet surface protein levels are more strongly associated with perioperative bleeding events than AVWS.

In conclusion, we demonstrated that VWF-related values including VWF-LMI, VWF:RCo/VWF:Ag, and maximal RIPA rates, but not expression levels of the platelet proteins GPIb or GPVI, could predict bleeding events in patients with LVADs.

### Limitations

A limitation of this study is that it was a retrospective single-center study analyzing a relatively small number of patients. Especially for platelet protein levels and RIPA, only 30 cases were analyzed.

## Conclusions

VWF-LMI, VWF:RCo/VWF:Ag, and maximal RIPA rates can be predictive factors of bleeding events in patients with LVADs.

## Data Availability

The datasets generated and/or analyzed during the current study are available from the corresponding author on reasonable request.

## References

1. McNamara N, Narroway H, Williams M, Brookes J, Farag J, Cistulli D, Bannon P, Marasco S, Potapov E, Loforte A. Contemporary outcomes of continuous-flow left ventricular assist devices-a systematic review. Ann Cardiothorac Surg. 2021;10:186–208. doi: 10.21037/acs-2021-cfmcs-35

2. Michelis KC, Zhong L, Peltz M, Pandey A, Tang WHW, Rohatgi A, Young JB, Drazner MH, Grodin JL. Dynamic Forecasts of Survival for Patients Living With Destination Left Ventricular Assist Devices: Insights From INTERMACS. J Am Heart Assoc. 2020;9:e016203. doi: 10.1161/JAHA.119.016203

3. Llerena-Velastegui J, Santafe-Abril G, Villacis-Lopez C, Hurtado-Alzate C, Placencia-Silva M, Santander-Aldean M, Trujillo-Delgado M, Freire-Ona X, Santander-Fuentes C, Velasquez-Campos J. Efficacy and Complication Profiles of Left Ventricular Assist Devices in Adult Heart Failure Management: A Systematic Review and Meta-analysis. Curr Probl Cardiol. 2023:102118. doi: 10.1016/j.cpcardiol.2023.102118

4. Matsumoto M, Kawaguchi S, Ishizashi H, Yagi H, Iida J, Sakaki T, Fujimura Y. Platelets treated with ticlopidine are less reactive to unusually large von Willebrand factor multimers than are those treated with aspirin under high shear stress. Pathophysiol Haemost Thromb. 2005;34:35–40. doi: 10.1159/000088546

5. Horiuchi H, Doman T, Kokame K, Saiki Y, Matsumoto M. Acquired von Willebrand Syndrome Associated with Cardiovascular Diseases. J Atheroscler Thromb. 2019;26:303–314. doi: 10.5551/jat.RV17031

6. Chen Z, Mondal NK, Ding J, Gao J, Griffith BP, Wu ZJ. Shear-induced platelet receptor shedding by non-physiological high shear stress with short exposure time: glycoprotein Ibalpha and glycoprotein VI. Thromb Res. 2015;135:692–698. doi: 10.1016/j.thromres.2015.01.030

7. Chen Z, Zhang J, Li T, Tran D, Griffith BP, Wu ZJ. The impact of shear stress on device-induced platelet hemostatic dysfunction relevant to thrombosis and bleeding in mechanically assisted circulation. Artif Organs. 2020;44:E201–E213. doi: 10.1111/aor.13609

8. Lukito P, Wong A, Jing J, Arthur JF, Marasco SF, Murphy DA, Bergin PJ, Shaw JA, Collecutt M, Andrews RK, et al. Mechanical circulatory support is associated with loss of platelet receptors glycoprotein Ibalpha and glycoprotein VI. J Thromb Haemost. 2016;14:2253–2260. doi: 10.1111/jth.13497

9. Mondal NK, Chen Z, Trivedi JR, Sorensen EN, Pham SM, Slaughter MS, Griffith BP, Wu ZJ. Association of Oxidative Stress and Platelet Receptor Glycoprotein GPIbalpha and GPVI Shedding During Nonsurgical Bleeding in Heart Failure Patients With Continuous-Flow Left Ventricular Assist Device Support. ASAIO J. 2018;64:462–471. doi: 10.1097/MAT.0000000000000680

10. Akiyama M, Sakatsume K, Sasaki K, Kawatsu S, Yoshioka I, Takahashi G, Kumagai K, Adachi O, Saiki Y. The incidence, risk factors, and outcomes of gastrointestinal bleeding in patients with a left ventricular assist device: a Japanese single-center cohort study. J Artif Organs. 2020;23:27–35. doi: 10.1007/s10047-019-01138-y

11. Bartoli CR, Zhang DM, Hennessy-Strahs S, Kang J, Restle DJ, Bermudez C, Atluri P, Acker MA. Clinical and In Vitro Evidence That Left Ventricular Assist Device-Induced von Willebrand Factor Degradation Alters Angiogenesis. Circ Heart Fail. 2018;11:e004638. doi: 10.1161/CIRCHEARTFAILURE.117.004638

12. Kang J, Hennessy-Strahs S, Kwiatkowski P, Bermudez CA, Acker MA, Atluri P, McConnell PI, Bartoli CR. Continuous-Flow LVAD Support Causes a Distinct Form of Intestinal Angiodysplasia. Circ Res. 2017;121:963–969. doi: 10.1161/CIRCRESAHA.117.310848

13. Uriel N, Colombo PC, Cleveland JC, Long JW, Salerno C, Goldstein DJ, Patel CB, Ewald GA, Tatooles AJ, Silvestry SC, et al. Hemocompatibility-Related Outcomes in the MOMENTUM 3 Trial at 6 Months: A Randomized Controlled Study of a Fully Magnetically Levitated Pump in Advanced Heart Failure. Circulation. 2017;135:2003–2012. doi: 10.1161/CIRCULATIONAHA.117.028303

14. Mehra MR, Goldstein DJ, Cleveland JC, Cowger JA, Hall S, Salerno CT, Naka Y, Horstmanshof D, Chuang J, Wang A, et al. Five-Year Outcomes in Patients With Fully Magnetically Levitated vs Axial-Flow Left Ventricular Assist Devices in the MOMENTUM 3 Randomized Trial. JAMA. 2022;328:1233–1242. doi: 10.1001/jama.2022.16197

15. Cai J, Xia W, Greenberg P, Okwuosa I, Setoguchi S, Akhabue E. Relation of Sociodemographic Factors With Primary Cause of Hospitalization Among Patients With Left Ventricular Assist Devices (from the National Inpatient Sample 2012 to 2017). Am J Cardiol. 2022;180:81–90. doi: 10.1016/j.amjcard.2022.06.047

16. Tamura T, Horiuchi H, Imai M, Tada T, Shiomi H, Kuroda M, Nishimura S, Takahashi Y, Yoshikawa Y, Tsujimura A, et al. Unexpectedly High Prevalence of Acquired von Willebrand Syndrome in Patients with Severe Aortic Stenosis as Evaluated with a Novel Large Multimer Index. J Atheroscler Thromb. 2015;22:1115–1123. doi: 10.5551/jat.30809

17. Sakatsume K, Akiyama M, Saito K, Kawamoto S, Horiuchi H, Saiki Y. Intractable bleeding tendency due to acquired von Willebrand syndrome after Jarvik 2000 implant. J Artif Organs. 2016;19:289–292. doi: 10.1007/s10047-016-0896-7

18. Sakatsume K, Saito K, Akiyama M, Sasaki K, Kawatsu S, Takahashi G, Adachi O, Kawamoto S, Horiuchi H, Saiki Y. Association between the severity of acquired von Willebrand syndrome and gastrointestinal bleeding after continuous-flow left ventricular assist device implantation. Eur J Cardiothorac Surg. 2018;54:841–846. doi: 10.1093/ejcts/ezy172

19. Tamura T, Horiuchi H, Obayashi Y, Fuki M, Imanaka M, Kuroda M, Nishimura S, Amano M, Sakamoto J, Tamaki Y, et al. Acquired von Willebrand syndrome in patients treated with veno-arterial extracorporeal membrane oxygenation. Cardiovasc Interv Ther. 2019;34:358–363. doi: 10.1007/s12928-019-00568-y

20. Shiraishi Y, Tachizaki Y, Inoue Y, Hayakawa M, Yamada A, Kayashima M, Matsumoto M, Horiuchi H, Yambe T. Hemolysis and von Willebrand factor degradation in mechanical shuttle shear flow tester. J Artif Organs. 2021;24:111–119. doi: 10.1007/s10047-020-01219-3

21. Sakatsume K, Akiyama M, Sakota D, Hijikata W, Horiuchi H, Maruyama O, Saiki Y. Individual Variability in von Willebrand Factor Fragility in Response to Shear Stress: A Possible Clue for Predicting Bleeding Risk. Asaio j. 2022;68:1128–1134. doi: 10.1097/mat.0000000000001623

22. Oishi H, Okada Y, Suzuki Y, Hirama T, Ejima Y, Fujimaki SI, Sugawara S, Okubo N, Horiuchi H. Impact of intraoperative use of venovenous extracorporeal membrane oxygenation on the status of von Willebrand factor large multimers during single lung transplantation. J Thorac Dis. 2023;15:4262–4272. doi: 10.21037/jtd-23-275

23. Yashige M, Inoue K, Zen K, Kobayashi R, Nakamura S, Fujimoto T, Takamatsu K, Sugino S, Yamano M, Yamano T, et al. Gastrointestinal Angiodysplasia before and after Treatment of Severe Aortic Stenosis. N Engl J Med. 2023;389:1530–1532. doi: 10.1056/NEJMc2306027

24. Okubo N, Sugawara S, Fujiwara T, Sakatsume K, Doman T, Yamashita M, Goto K, Tateishi M, Suzuki M, Shirakawa R, et al. VWF:RCo/VWF:Ag for diagnosis of acquired von Willebrand syndrome caused by aortic stenosis. Research and Practice in Thrombosis and Haemostasis. 2023. doi: 10.1016/j.rpth.2023.102284

25. de Jong A, Dirven RJ, Oud JA, Tio D, van Vlijmen BJM, Eikenboom J. Correction of a dominant-negative von Willebrand factor multimerization defect by small interfering RNA-mediated allele-specific inhibition of mutant von Willebrand factor. J Thromb Haemost. 2018;16:1357–1368. doi: 10.1111/jth.14140

26. Boender J, Atiq F, Cnossen MH, van der Bom JG, Fijnvandraat K, de Meris J, de Maat MPM, van Galen KPM, Laros-van Gorkom BAP, Meijer K, et al. Von Willebrand Factor Multimer Densitometric Analysis: Validation of the Clinical Accuracy and Clinical Implications in Von Willebrand Disease. Hemasphere. 2021;5:e542. doi: 10.1097/HS9.0000000000000542

27. Nakamura M, Imamura T, Ueno H, Kinugawa K. Impact of the Severity of Acquired von Willebrand Syndrome on the Short-Term Prognosis in Patients with Temporary Mechanical Circulatory Support. Medicina (Kaunas). 2022;58. doi: 10.3390/medicina58020238

28. Pienta MJ, Wu X, Cascino TM, Brescia AA, Abou El Ela A, Zhang M, McCullough JS, Shore S, Aaronson KD, Thompson MP, et al. Advancing Quality Metrics for Durable Left Ventricular Assist Device Implant: Analysis of the Society of Thoracic Surgeons Intermacs Database. Ann Thorac Surg. 2022;113:1544–1551. doi: 10.1016/j.athoracsur.2022.01.031

29. Yaoita N, Shirakawa R, Fukumoto Y, Sugimura K, Miyata S, Miura Y, Nochioka K, Miura M, Tatebe S, Aoki T, et al. Platelets are highly activated in patients of chronic thromboembolic pulmonary hypertension. Arterioscler Thromb Vasc Biol. 2014;34:2486–2494. doi: 10.1161/ATVBAHA.114.304404

30. James PD, Connell NT, Ameer B, Di Paola J, Eikenboom J, Giraud N, Haberichter S, Jacobs-Pratt V, Konkle B, McLintock C, et al. ASH ISTH NHF WFH 2021 guidelines on the diagnosis of von Willebrand disease. Blood Adv. 2021;5:280–300. doi: 10.1182/bloodadvances.2020003265

